# Pre-frontal cortical activity during gait is altered in pre-manifest and early spinocerebellar ataxia

**DOI:** 10.1101/2024.11.14.24317186

**Authors:** Martina Mancini, Carla Silva-Batista, Vrutangkumar V. Shah, Fay B. Horak, Patricia Carlson-Kuhta, Delaram Safarpour, Christopher M. Gomez

**Affiliations:** Department of Neurology, Oregon Health & Science University, Portland, Oregon, USA; APDM Precision Motion, Clario, Portland, Oregon, USA; Department of Neurology, The University of Chicago, Chicago, Illinois, USA

## Abstract

**Background:** Spinocerebellar ataxia (SCA) is a degenerative cerebellar disease, causing progressive impairment of gait and balance in adults. To identify the ideal subjects for disease-modifying therapies it is critical to identify biomarkers for the earliest stages of SCA.

**Objective:** We investigated whether prefrontal cortex activity is increased during walking in in early SCA or in pre-manifest SCA compared to healthy control subjects.

**Methods:** Sixteen participants with genetically determined SCA and 15 age-matched healthy controls participated in the study. The SARA was administered by a movement disorders specialist before the gait assessment. An 8-channel, mobile, fNIRS, with two reference channels, was used to record changes in oxygenated hemoglobin (HbO_2_) and deoxygenated hemoglobin within the PFC. Participants walked for 2-minutes at a comfortable pace while wearing wireless, inertial sensors to derive gait characteristics.

**Results:** Of the 16 individuals with SCA, 9 were classified as pre-manifest (SARA<3) and 7 as early SCA (SARA<10). PFC activity (HbO_2_) while walking was greater than controls of similar age in people with SCA. Increased PFC activity was also present even in the pre-manifest stage of SCA. Increase in PFC activity was related to worse gait (double-support time and toe-out angle).

**Conclusions:** PFC activity is increased in pre-manifest SCA, even when clinical scores are normal in the pre-manifest stage of the disease, and may serve as a biomarker that precedes onset of clinical disease. Increased PFC activity is consistent less automatic, cortical control of gait to compensate for impaired automatic, cerebellar control, even in early stages of ataxia.

## Introduction

The autosomal-dominant SCAs are a group of neurodegenerative diseases characterized by progressive impairment of gait and balance, as well as dysarthria and limb incoordination^1, 2^. The clinical symptoms in the SCAs usually begin in adulthood and the degenerative process involves the cerebellar cortex and, variably, other brain regions, depending on the genetic subtype^3-5^.

Recent advances in preclinical studies of the SCA^6-8^ have raised the promise of targeted, disease-modifying therapies designed to stop or slow disease progression. In animal models, elimination of the offending, ataxia gene results in improvement of function and neuroprotection, particularly in the early or pre-manifest stages of the disease model^6-8^. Clinical trials for these rare neurodegenerative diseases are hampered by: 1) the inadequacy of clinical outcome assessments, that provide only semiquantitative assessments of disease progression and 2) incomplete understanding of the pre-manifest stages of SCA^6, 9^. Although numerous pharmaceutical companies are in the planning stages for Phase 1 and 2 clinical trials of gene therapies, there is growing recognition of the need for early biomarkers of SCA to empower clinical trials.

SCA is characterized by a prolonged pre-manifest stage, defined as the interval from birth to the earliest clinical appearance of cerebellar deficits. This interval is of great interest because this stage could provide a window for early therapeutic intervention before irreversible neuronal damage has occurred^6-8^. Preclinical studies in genetic mouse models in which the ataxia-causing transgene can be switched off at different times suggest that the ability to treat the SCAs by modulating the underlying cause will wane as the disease progresses. Early reversibility suggests that functional alterations precede irreversible structural changes^10^.

The four most common SCAs, SCA 1, 2, 3, and 6, are fully penetrant, yet despite some non-motor symptoms, healthy carriers of these mutations commonly experience a long disease-free, pre-manifest period. When studied with sensitive performance tests patients, at least late in the pre-manifest period, some patients may manifest various functional abnormalities such as ocular-motor changes and increased variability of some balance and gait parameters^11-16^. Moreover, subtle structural and neurochemical changes in the cerebellum and cerebral cortex can be detected prior to the onset of clinically detectable balance and gait abnormalities in SCA, particularly abnormalities in variability of gait^17-19^. These observations suggest that, in the period before onset of overt clinical changes, sensitive measures may detect evidence of the disease process.

From a physiological standpoint, gait is characterized by functioning of lower, spinal and brainstem circuitry in a process termed “automaticity of gait”, requiring limited prefrontal cortical control^20, 21^. In normal subjects at the beginning of a walking task there is brief, initial increase in prefrontal cortex activity, followed by a decline in PFC activity, in a process referred to as a “return to automaticity”^22^. These results suggest a return to an automatic control of gait following an initial attentional period at gait initiation. Because of well-documented functional and anatomic connections between the frontal lobe and the cerebellum, it has been hypothesized that increased prefrontal cortex (PFC) activity during gait in SCA might provide a critical compensatory role in the presence of impaired cerebellar control of gait and balance^23-25^.

Recent development of wireless, functional near infrared spectroscopy (fNIRS) of the brain provides direct, physiological measures of automaticity, such as PFC activity while walking. Although this technique is nowadays widely used in investigating cortical correlates of gait in neurological diseases, only 2 studies have reported PFC activity during gait in SCAs and none have examined PFC activity in pre-symptomatic SCAs^23, 24^. Understanding the role of the pre-frontal cortex while walking may help in predicting timing of symptoms onset, and its potential role in disease modulation and designing therapies. Here, our objective was to investigate the PFC activity while walking in a group of people with genetically confirmed SCA that do not show any clinical symptoms (pre-manifest SCA), and a group who shows mild symptoms (early SCA). We hypothesize that loss of gait automaticity in clinically manifest SCA may be associated with increased PFC activity, and that this increased PFC activity may appear in the premanifest period, well before the onset of clinical changes. We further hypothesize that SCA patients in both the manifest and premanifest period will sustain increased PFC activity throughout the walking task, demonstrating a failure to return to gait automaticity.

## Methods

### Participants

Sixteen people with genetically confirmed SCAs and 15 healthy controls of similar age were included in the study. Participants were included if they had a positive gene test for SCA and a SARA score of 0-9 and ability to stand for 30 seconds and walk independently for 2 minutes. Subjects were divided into those with SARA scores < 3 termed, pre-manifest, and 3-9 termed early manifest ataxia. Exclusion criteria included: factors affecting gait (e.g., musculoskeletal disorders, uncorrected vision or vestibular problems) and inability to follow instructions. Study procedures were approved by the University Institutional Review Board (eIRB #9903), with written informed consent obtained prior to participation.

### Experimental procedures and equipment

Participant characteristics of age, sex, and disease duration were recorded. Disease severity was measured using the SARA^26^ administered by a movement disorders specialist.

Participants walked, at self-selected, comfortable pace, back and forth over a 10 m straight path, with a 180° turn at each end. Two conditions were tested: single and dual-task walking. The cognitive dual-task was added to challenge the gait performance in these early stage individuals. Each condition included an initial 20 s of quiet standing (baseline period) followed by 80 s of walking (task period). The dual-task condition consisted of executing the walking task while performing a concurrent, cognitive task (auditory Modified AX-Continuous Performance Task),^27^ which required participants to press a handheld button after a two-paired letters sequence. The sequence consisted of a cue letter ‘‘A” and a probe letter ‘‘I” presented sequentially so that the target trail was ‘‘AI” and participants were asked to respond as fast as possible after the probe letter. No information about task priority was assigned to participants not to influence the task execution. A research assistant walked with the participants to avoid falls and ensure their safety.

A portable, 8-channel fNIRS system (OctaMon, Artinis Medical Systems, Elst, The Netherlands) recorded changes in HbO_2_ and HHb (oxygenated and deoxygenated hemoglobin, respectively) bilaterally in the PFC at a sampling rate of 50 Hz. The fNIRS device consisted of two light detectors and eight light emitters (continuous wave diodes with wavelengths of 760 and 850 nm). Three regular channels (interoptode distance of 35 mm) and one short-separation channel (interoptode distance 15 mm) were used for each hemisphere. Optodes were placed on participants forehead using a headband with predetermined locations (according to the international 10-20 EEG system). A digitizer (Polhemus Patriot 3D digitizer, Colchester, VT, USA) was used to provide 3-dimensional coordinates of anatomical references (Cz, nasion, inion and left and right preauricular points) and positions of optodes.

Six inertial measurement units (Opals by APDM-Clario) were used to quantify spatiotemporal gait parameters at a sampling rate of 128 Hz. Opals were located on the sternum and pelvis, on the wrists, and both feet of participants. Each inertial sensor consisted of tri-axial accelerometers, gyroscopes, and magnetometers, and were securely fixed to the participant’s body with Velcro straps.

### Data analysis

Processing of fNIRS signals followed previously published recommendations^28, 29^. The spatial registration routine was used to find the correspondence between the scalp location where the fNIRS measurement was performed and its underlying cortical surface where the source signal was located^30^. Cortical regions assessed included the Brodmann areas 9 and 10 over the prefrontal cortex.

The fNIRS data were preprocessed within custom-made MATLAB algorithms, which consisted of several steps. Specifically: 1) raw intensity data conversion into optical density, 2) artifacts correction by wavelet filtering;^31^ 3) optical density data conversion to HbO_2_ and HHb concentrations; 4) correlation-based signal improvement method to attenuate remaining artifacts.^32^ The next step involved removing superficial hemodynamic response from the long-distance channels (3.5cm), using the short-distance channels (1.5cm)^33^. Briefly, scaling factors were determined by detecting the peaks (positive and negative) of the heart rate within the long and short-separation channel signals, then dividing them to produce the scaling factors for each pair of channels. A low-pass filter with a cut-off frequency of 0.14 Hz removed any remaining high-frequency noise and signals in the 6 long-distance channels were median-averaged. The data were then baseline-adjusted by subtracting the mean of the baseline period (standing) from the entire trial. Lastly, relative changes in HbO_2_ and HHb concentrations were calculated for both early (median of the first half of the task = initial 40 s) and late (median of the second half of the task = final 40 s) phases of the walking task.

Spatiotemporal gait measures were calculated from the inertial sensors using the *Mobility Lab* software, *V2* (APDM-Clario).^34^ All recorded steps corresponding to walking were included in the analysis. The following gait measures were reported accordingly to our previous paper, as most sensitive lower limb measures in early stage SCA and related to dynamic postural instability while walking^11^: 1) toe-out angle variability, 2) double-support time variability, 3) average toe-off angle, 4) toe-off angle variability, and 5) elevation at mid-swing variability. In addition, we reported average gait speed and double-support time, as commonly used in this population^35^.

### Statistical Analysis

Non-parametric tests were used to compare differences between groups. To compare PFC activity and gait measures across the three groups, the Kruskal-Wallis test was used. When the Kruskal-Wallis test yielded a significant effect (p < 0.05), post-hoc analysis was performed using a Wilcoxon Rank Sum test. To compare changes in PFC activity while walking between single-task and dual-task, the Wilcoxon signed-rank test was used. Lastly, to investigate the association between PFC activity and gait measures we used Spearman’s rank correlations. All statistical analysis was performed using MATLAB 2021b (The Mathworks Inc., Natick, MA, USA). The statistical significance was set to P < 0.05.

## Results

Of the 16 individuals with SCA, 9 were classified as pre-manifest (SARA<3) and 7 as early manifest SCA (SARA<10) with an age range of 33 – 70 years old, same for the 15 healthy controls subjects.

Gait measures are reported in Table 1. Aspects of gait variability, such as toe-out angle, double-support time, and elevation at mid-swing were significantly different across groups (p-values varying from 0.01 and 0.003). However, post-hoc tests indicated that group differences in gait variability were only present between healthy controls and people with early manifest SCA, as well as between pre-manifest versus early manifest, but not between healthy controls and pre-manifest SCA (Table 1). The average gait speed, double-support time, toe-off angle, and toe-off angle variability were similar across the three groups (p>0.05).

**Table 1.** Group differences for the gait characteristics.

|  | Healthy controls (HC) |  | Pre-manifest SCA (PRE) |  | Early manifest SCA (EARLY) |  | Statistics |  |  |  |  |
| --- | --- | --- | --- | --- | --- | --- | --- | --- | --- | --- | --- |
|  | Mean | Std | Mean | Std | Mean | Std | Chi-square | P-value | P (HC vs PRE) | P (HC vs EARLY) | P (PRE vs EARLY) |
| Gait speed (m/s) | 1.16 | 0.19 | 1.19 | 0.13 | 1.12 | 0.19 | 0.74 | 0.8 |  |  |  |
| Double-support time (% gait cycle) | 21.06 | 2.8 | 20.4 | 1.8 | 20.4 | 3.9 | 0.22 | 0.8 |  |  |  |
| Double support time variability (% gait cycle) | 1.25 | 0.35 | 1.35 | 0.25 | 2.02 | 0.51 | 11.2 | 0.003 | 0.22 | 0.003 | 0.003 |
| Toe-off angle (degrees) | 36.5 | 5.4 | 36.5 | 4.8 | 32 | 4.3 | 4 | 0.13 |  |  |  |
| Toe-off angle variability (degrees) | 1.6 | 0.7 | 1.6 | 0.5 | 2.1 | 0.6 | 3.85 | 0.14 |  |  |  |
| Toe-out angle variability (degrees) | 2.8 | 0.6 | 3.1 | 0.7 | 4 | 1 | 6.11 | 0.04 | 0.48 | 0.01 | 0.09 |
| Elevation at mid-swing variability (degrees) | 0.35 | 0.19 | 0.41 | 0.07 | 0.59 | 0.17 | 8.67 | 0.01 | 0.16 | 0.01 | 0.02 |

### Both Pre-manifest and Manifest SCA show increased PFC activity during walking compared to healthy controls

Average PFC activity (HbO2) while walking was significantly different between healthy controls and people with SCA, see Figure 1 (Chi-square=15.66, p=0.0004). Specifically, we observed an increase in PFC activity in pre-manifest SCA as well as early manifest SCA compared to healthy controls (p=0.001 and p=0.002, respectively). PFC activity while walking was similar in pre-manifest versus early SCA (P=0.84).

**Figure 1.**
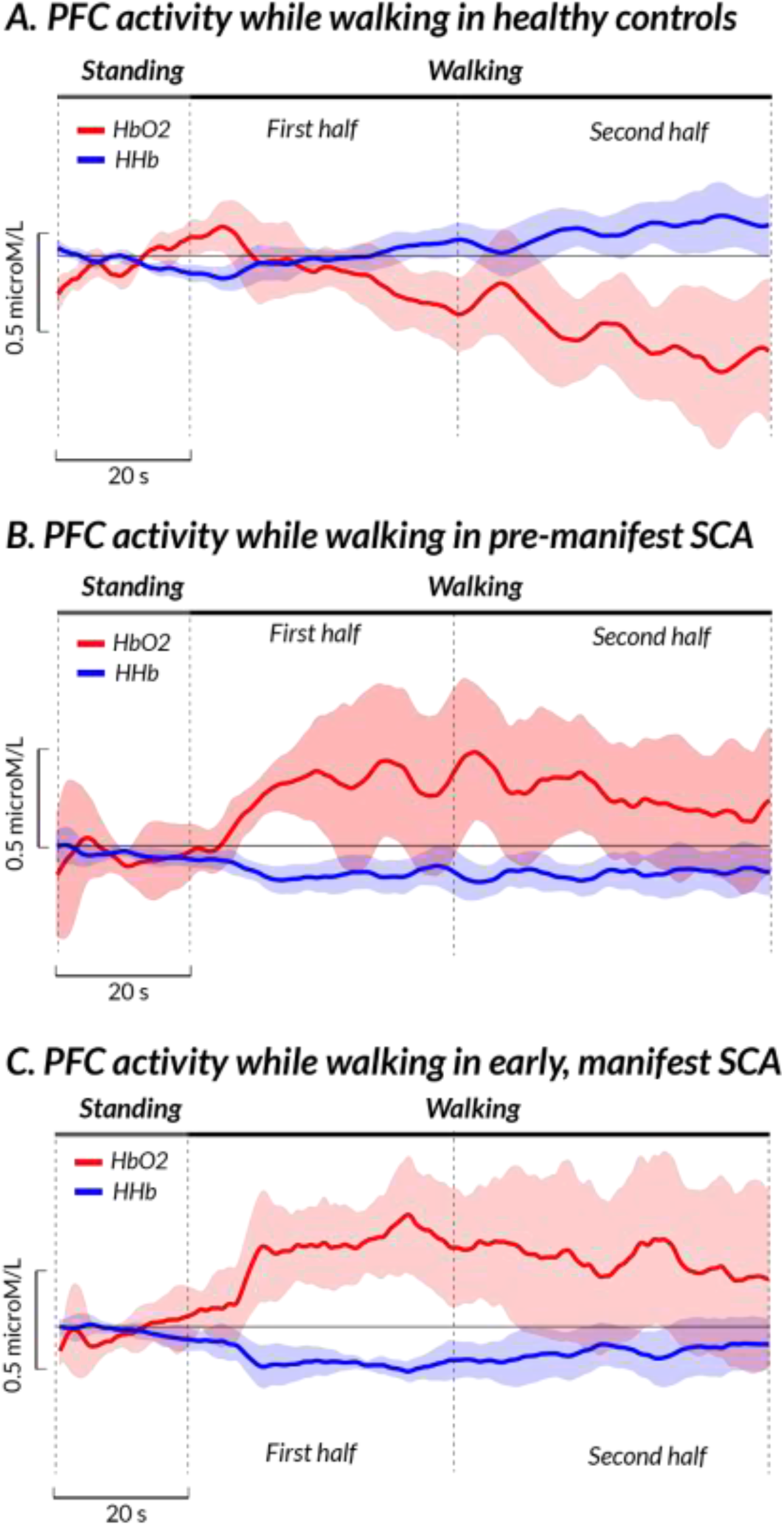
Boxplot with median, 25^th^ and 75^th^ percentile (and individual subjects) for the relative HbO2 while walking at the PFC for the 3 groups. HbO2: oxygenated hemoglobin.

When adding a concurrent dual-task to walking, PFC activity was increased compared to single task walking only in the early manifest SCA group (p=0.04) but not in the pre-manifest SCA group (p=0.82).

### Both pre-manifest and early manifest SCA have sustained PFC activity throughout the walking

As expected, the healthy control group showed a significant decrease in PFC activity in the second minute of the walking task compared to the first minute (P<0.001), Figure 2. However, neither the pre-manifest nor early manifest SCA significantly decreased PFC over the course of the 2-minute walk task (Figure 2).

**Figure 2.**
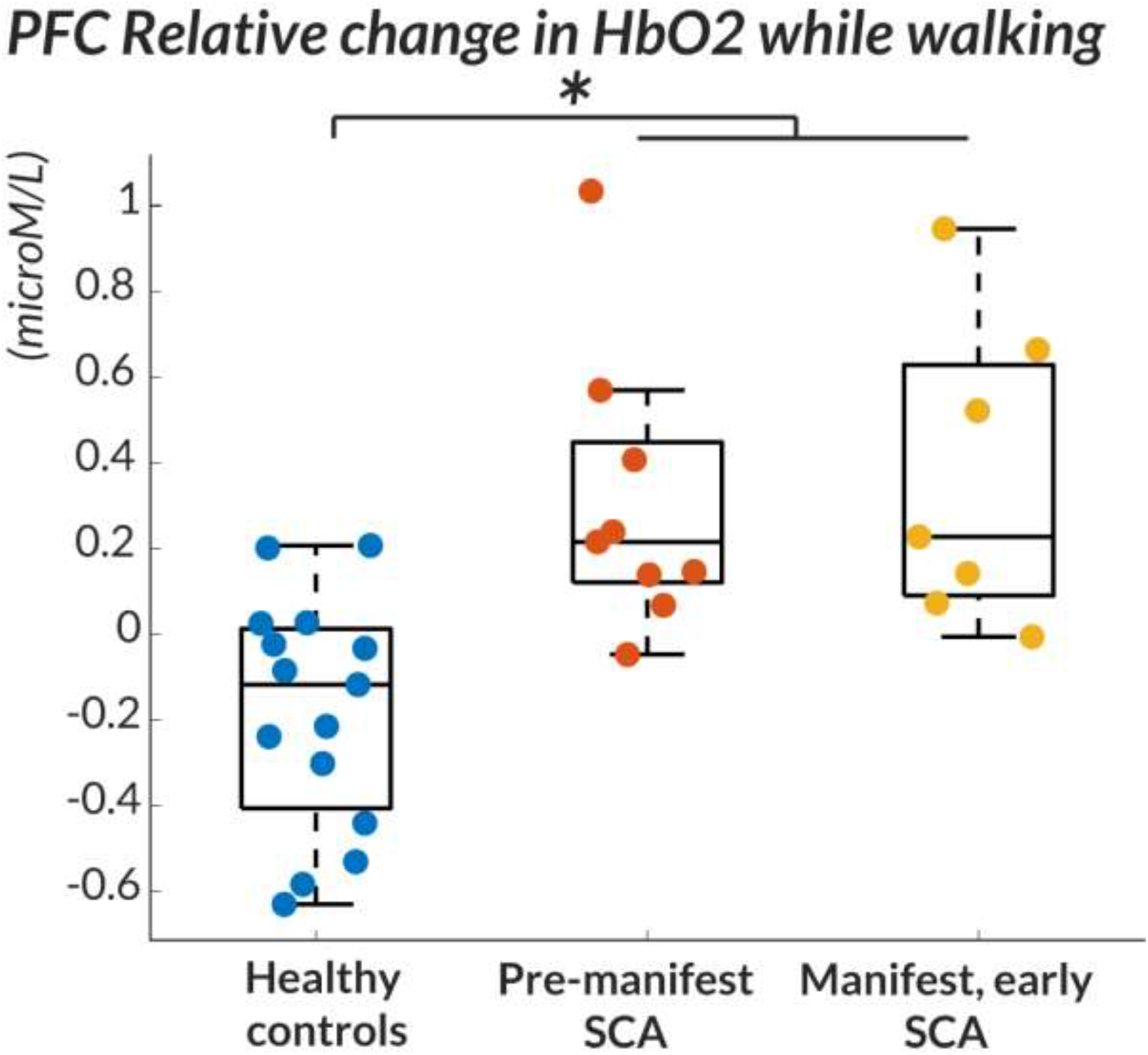
Group average of HbO2 (red bold line) and HHb (blue bold line) signals during 20-seconds standing still and 80-seconds walking task in healthy controls (A), people with pre-manifest SCA(B) and people with early SCA(C). The shadow is 95% confidence interval.

### Greater PFC activity in pre-manifest and early manifest SCA is associated with greater gait impairments

When looking at the pre-manifest and early SCA data together, we observed a significant correlation between increase in PFC activity while walking and increased double-support time (r=0.52, p=0.03), Figure 3. No correlations were present between PFC activity and other gait spatio-temporal parameters nor between PFC activity during gait and clinical SARA scores.

**Figure 3.**
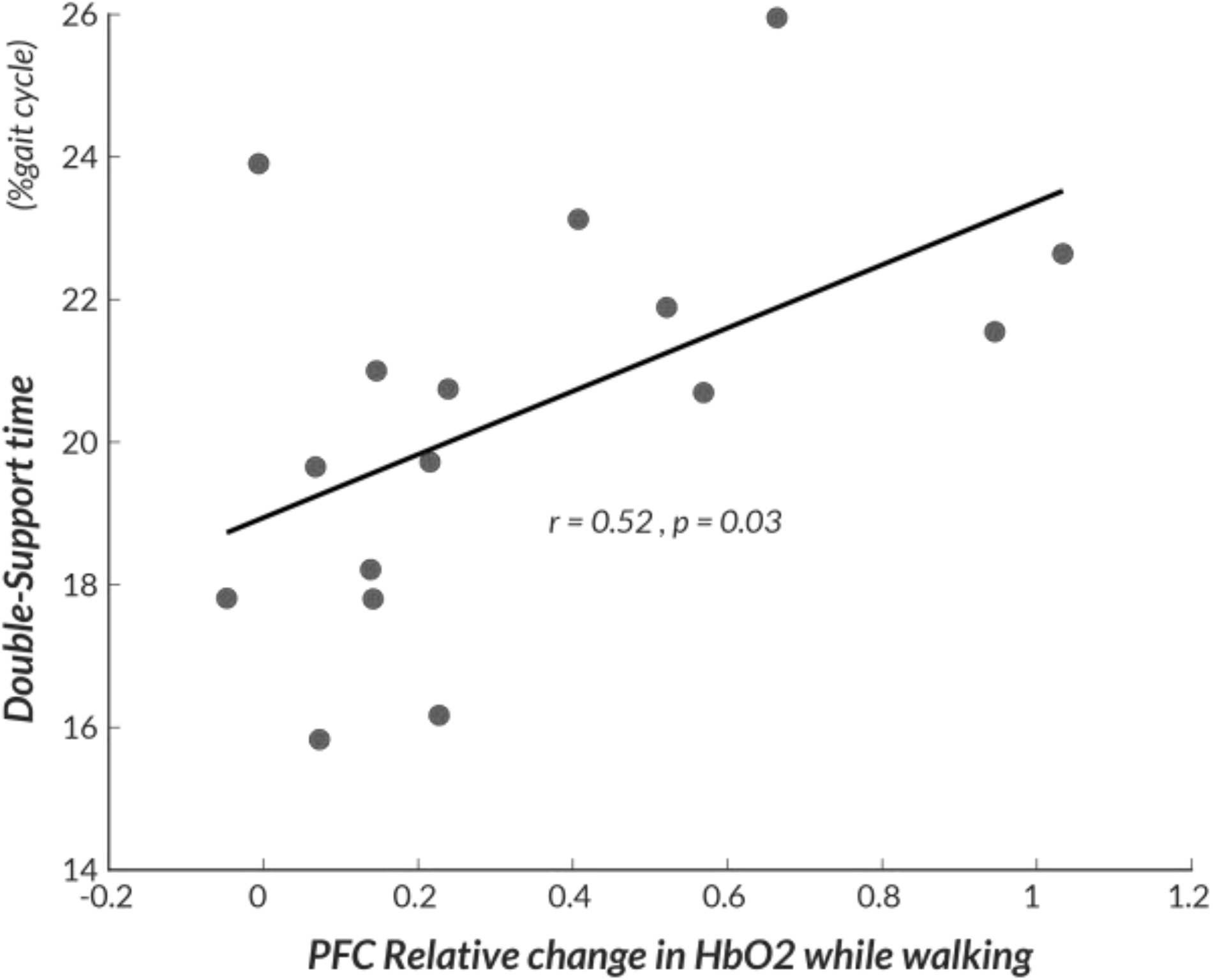
Scatterplot of the correlation between PFC activity while walking and double support time in the participants with SCA.

## Discussion

To our knowledge, this is the first study assessing the feasibility of measuring abnormal PFC activity while walking in people with pre-manifest and early manifest SCA. Consistent with our hypothesis, our pilot findings suggest that PFC activity while walking is increased, not only in early SCA, but also in pre-manifest SCA, when quantitative gait characteristics are similar to age-matched healthy controls. The results indicate that increased PFC activity during walking may appear prior to the onset of objective signs of gait impairment in genetically at-risk SCA patients. While these findings provide insight into motor control and the disease process, this also may have important implications for defining a critical prodromal period prior to the onset of permanent neurological dysfunction in the context of potential neuroprotective therapies.

Increased PFC activity is consistent with requirement for greater cortical control of gait to compensate for impaired automatic, cerebellar control^36^. Increased PFC activity in early manifest SCA, as with other gait disorders, may reflect compensation for cerebellar dysfunction^23, 24^ and suggests impairment of gait automaticity. In pre-manifest SCA, in the absence of detectable gait impairment, it is possible that PFC activity while walking may increase unconsciously to maintain a stable walking behavior. In fact, our results showed that both pre-manifest and early SCA had a sustained increase in PFC activity throughout the walking task while healthy controls displayed a return to automatic gait control following an initial attentional period at gait initiation.

The cerebellum is a quick, automatic, sensorimotor (proprioceptive, vestibular and visual) feedback system that is thought to compare intended action with actual performance and then correct the subsequent movements^37, 38^. Thus, for example, if a step has been found to be wider (larger toe out angle) or double-support time longer (impaired balance) than intended, the cerebellum’s role is to improve the next steps. Increased movement (and step by step) variability is, thus, a hallmark of cerebellar dysfunction, as less automatic, longer duration sensorimotor cortical pathways, attempt to correct stepping movement errors^39, 40^. It is possible that people with pre-manifest SCA increase their PFC activity while walking to achieve stable walking behavior, thus implicating that changes in PFC activity may precede changes in gait. Although we did not observe any significant change in variability of toe-out angle, double-support time, nor foot elevation at mid-swing between pre-manifest SCA and healthy controls, this may be due to our small sample size as previous studies^11, 41^ have identified gait variability measures that were already altered in pre-manifest SCA. Nevertheless, the increase in PFC activity was robust and significant.

As gait variability is thought to be a surrogate measure of reduced locomotor automaticity (more PFC control), the onset of excessive PFC activity may be a more sensitive early marker of failing cerebellar function than quantitative gait variability measures in pre-manifest SCA. Increased gait variability reflects abnormal dynamic balance control and impaired gait stability (equilibrium control) while walking in cerebellar ataxia^38, 42 43-46^. Gait variability becomes increasingly impaired with progression of ataxia, although PFC activity may plateau early in pre-manifest SCA. The magnitude PFC activity did not correlate with the changes in several gait parameters, suggesting that PFC activity was maximally active, even with milder gait abnormalities. On the other hand, the magnitude of PFC activity did correlate with double-support time of gait, a sensitive measure of balance control while walking^47, 48^. Double-support time tends to increase with more severe ataxia, because of the difficulty controlling balance on one foot^35^. Although most of our small group of pre-manifest and early SCA did not yet show abnormally long double support time, this measure was positively correlated with PFC activity (Figure 3).

Identification of cortical compensatory mechanisms with novel, accurate, wearable technology will provide quantifiable and objective outcomes that could be used to predict ataxia symptom onset, to refine the therapeutic window or as targets for new therapies. Specifically, these cortical markers could be used to test the effectiveness of new, disease-modifying, gene therapies for SCA. Studies in larger populations and test-retest reliability are needed to confirm these promising pilot findings.

This study has several limitations, mainly due to a small sample size and cross-sectional nature of the study. Given that studies were conducted in SCA mutation carriers in whom the predicted age on onset was within 10 years, it remains to be determined whether the increases in PFC activity while walking are present during the lifetime of the genetically at-risk patients or whether the PFC activity during walking represents the stage immediately prior to the onset of gait abnormalities. Future work will need to confirm these findings in a larger cohort followed longitudinally to investigate whether PFC activity while walking could be a useful marker to predict symptoms onset in SCA.

## Data Availability

All data produced in the present study are available upon reasonable request to the authors

## Acknowledgment

The authors thank all participants for generously donating their time to participate and research assistants for scheduling data collection.

## Author Roles

MM: Study concept/design; Drafting/revising the manuscript; Data acquisition; Data analysis/interpretation; Writing the first manuscript draft.

V.V.S.: Revising the manuscript; Data interpretation.

C.S-B.: Revising the manuscript; Data acquisition; Data interpretation.

P.C.K.: Revising the manuscript; Data acquisition; Data interpretation.

F.B.H.: Drafting/revising the manuscript; Data interpretation; Study concept/design.

D.S.: Revising the manuscript; Data interpretation.

C.M.G.: Drafting/revising the manuscript; Data interpretation; Data acquisition; Study concept/design.

